# Does performance on the Ages and Stages Questionnaire-3 assessment at age 2 estimate the effect on the Early Years Foundation Stage at age 5? A longitudinal observational study using routine data

**DOI:** 10.64898/2026.02.27.26347090

**Authors:** J Dickerson, Y Xu, R Shore, H.C. Henderson, D Lee, K Bennett, P Degnan, K Sohal, M Mon Williams, J Wright, K.E. Mooney

## Abstract

**Introduction:** Improving the proportion of children achieving a good level of development (GLD) on the Early Years Foundation Stage Profile (EYFSP) aged five became a national priority in England in 2025. This study examined whether performance on the Ages and Stages Questionnaire-3 (ASQ-3) aged two can estimate the effect on the EYFSP.

**Methods:** A longitudinal dataset was created using linked Connected Yorkshire (Bradford) data. Multiple regression analyses estimated the effect of ASQ-3 on EYFSP and descriptive analyses explored the overlap between GLDs.

**Results:** Of 98,851 children born between September 2013 to May 2023, 47,046 (48%) had an ASQ-3 record and 6,021 had linked EYFSP data. Children who achieved a GLD on the ASQ-3 had over three-times the odds of achieving a GLD on the EYFSP (OR 3.18, 95% CI 2.70-3.75). Children who do not achieve a GLD on the ASQ-3 are very likely to not achieve a GLD on the EYFSP, however the ASQ-3 misses identification of 3 in 4 children who will not achieve a GLD on the EYFSP. Boys, those living in areas of deprivation, and those from White Other backgrounds were less likely to achieve an EYFSP GLD.

**Conclusion:** This study highlights the value of using ASQ-3 to identify children who will not achieve a GLD on the EYFSP. However, the ASQ-3 misses many other children at risk of not achieving a GLD on the EYFSP. There are inequities in early child development, and early identification and effective intervention are critical to reduce these inequities.

**Key Messages:** 

**What is already known on this topic:**

□ In England, approximately 3 in 10 children do not reach a Good Level of Development (GLD) on the Early Years Foundation Stage Profile (EYFSP) which is undertaken at the end of the Reception year of school. Reaching a GLD is associated with later educational attainment.

At age two, all children in England are offered a developmental assessment using the validated Ages and Stages Questionnaire (ASQ-3). The ASQ-3 was developed in the USA and adapted to British English.

**What this study adds:**

□ This study is the first to use longitudinal data to assess whether performance on the ASQ-3 estimates the effect on the EYFSP at school
□ This study found that children who did achieve a GLD on the ASQ-3 had more than three times the odds of achieving a GLD on the EYFSP. Children who do not achieve a GLD on the ASQ-3 are highly likely to also not achieve a GLD on the EYFSP, however the ASQ-3 misses identification of a high proportion of children who will go on to not achieve a GLD on the EYFSP.
□ Boys, those living in areas of high deprivation, and those from White Other backgrounds were less likely.

**How this study might affect research, practice or policy:**

□ This study highlights the value of using the ASQ-3 assessment to identify children who are not going to achieve a GLD on the EYFSP. However, the ASQ-3 only detects 1 in 4 children who will not be school ready, meaning that other methods are also needed to identify children at risk of poor school readiness.
□ these findings show that more than 1 in 10 children have identifiable developmental delay by two years of age, and that this is more likely in boys and those from sociodemographic disadvantage, emphasising the importance of intervening from the earliest possible moment to reduce inequities in child development.

## Introduction

As part of the UK Government’s drive to reduce inequalities in children’s development and longer-term life chances, it has set an ambitious target that by 2028, 75% of children in England should reach a Good Level of Development (GLD) at the end of their first year of schooling at age five. To support this, a new “Best Start in Life” programme will be rolled out nationally with each local authority required to roll out a local plan to improve school readiness of children in their area [1].

In England, school readiness is assessed by teachers using the Early Years Foundation Stage Profile (EYFSP) [2]. To achieve an overall ‘Good Level of Development’ (GLD) children need to achieve five core domains (communication and language, physical development, personal, social and emotional development, literacy and mathematics). A GLD on the EYFSP is a strong early indicator of educational success and developmental outcomes, [3-8].

Several sociodemographic characteristics are associated with not achieving a GLD, including male sex, children experiencing socioeconomic disadvantage, some ethnic minority groups, those with English as an additional language and children born prematurely [9-11].

While there is considerable evidence about the predictive utility of the EYFSP, the relationship between earlier developmental measures and the EYFSP is less well described. In England, all children are offered a two-year developmental review wherein development is assessed using the parent-reported Ages and Stages Questionnaire 3 (ASQ-3). Children achieve a GLD if they score are above a specified cut-off across five domains: gross and fine motor skills, personal-social development, communication and problem solving [12]. The ASQ-3 assessment in England is currently used as a population level measurement due to concerns about the lack of validation in England [13].

A clear understanding of whether performance on the ASQ-3 estimates the effect on school readiness could provide an opportunity to identify children who would benefit from early interventions and increase the number of children achieving a GLD on the EYFSP.

An area level, cross-sectional data analysis in England found that the percentage of children achieving a GLD on the ASQ-3 correlated weakly with the percentage of children achieving a GLD in the EYFSP [14]. However, the use of area level, cross-sectional data limits the validity of these results as it compares two different groups of children and relationships at an aggregated level. Furthermore, this analysis did not control for known sociodemographic predictors of ASQ-3 or EYFSP.

To date, no study has explored the estimated effect of ASQ-3 on EYFSP using longitudinal individual-level child data. One of the key barriers is that data from the 0-5 Healthy Child Programme is not linked to education and nursery data due to different organisations holding the data and using different identifiers (NHS and Unique Pupil Numbers).

This study bridges this gap by using the Connected Yorkshire (Bradford) (CYB) dataset -of routinely collected health, education and social care data linked using pseudonymised identifiers [15]. The aim of this study is to: a) describe the sociodemographic patterns of children achieving a GLD on the ASQ-3 and the EYFSP; and b) test whether children’s performance on the ASQ-3 aged two estimates the effect on their school readiness measured using the EYFSP aged five.

## Methods

### Study design

This is an observational study using individual longitudinal data and follows the STROBE checklist.

### Setting

Bradford is a district in the North of England which ranges from severely disadvantaged, ethnically diverse inner-city areas, to areas of low deprivation and lesser diversity in more suburban and rural areas [16]. Across the District 59% (∼4,000 per year) of children are born into the lowest decile of deprivation on the Index of Multiple Deprivation (IMD) [17]. In 2023-2024 the proportion of children achieving a GLD on the EYFSP was 62% compared to the national average of 69%, but varied from 47% in disadvantaged areas to 89% in areas of low deprivation [17].

CYB has ethical approval for the use of linked data for research purposes (Yorkshire & Humber - Bradford Leeds REC 18/YH/0200; East Midlands - Derby REC 22/EM/0127). The ‘base’ of CYB are primary care records from General Practices located within the Bradford District. A total of 323,793 primary care records between October 1999 and August 2025 are in the dataset and have been linked to secondary and community care, education and other services [15].

This study used primary care data linked to community care health visitor records for the ASQ-3, Department for Education data for the EYFS and school Census-level data to obtain ethnicity, sex, birth and death records and Lower Super Output Area (LSOA) to derive IMD. The ASQ-3 was introduced nationally on the 1st April 2015, with full implementation from 1^st^ September 2015 onwards, and is therefore available for all children born after September 2013. The education dataset was available up to and including the academic year 2018-2019 and is therefore available for children born from 1^st^ September 2013 to 31^st^ August 2014 (one full academic year) in this study.

### Sample Eligibility

All children in the CYB baseline dataset (registered with a general practice in the Bradford district) with a date of birth between September 2013 and May 2023 who met the following criteria:

□ ASQ-3 analyses: had an ASQ-3 assessment completed between 24 and 30 months of age, available from September 2015 to May 2025.
□ EYFSP analyses: had an EYFSP assessment recorded at the end of the school reception year for the academic year 2018-2019
□ ASQ-3 to EYFSP analyses: All children who had an ASQ-3 assessment between 24 to 30 months of age and had an EYFSP assessment recorded in the academic year 2018-2019.

### Patient and public involvement

This Study was co-designed with key stakeholders in Bradford, with input from national policy makers. As it uses secondary data analysis, there has been no public involvement specific to this study.

### Outcome measures

The ASQ-3 was derived into two formats:

A binary (GLD) indicator using the Public Health England definition (used for national statistics) [13,18]: GLD not achieved - any of the five domains (communication, gross motor, fine motor, personal-social, problem solving) classified as “Below Cut Off”; GLD achieved - all domains were either “No Risk” or “Monitor”.

A continuous ASQ-3 composite score summing the binary scores (1 = No Risk, 0 = Monitor/Below Cut Off) across all five domains, resulting in a score from 0 to 5, where higher values indicate lower developmental concern.

The EYFSP was derived into two formats:

A binary GLD variable using national criteria [19]: GLD achieved if they meet the expected level on the five core domains.

A continuous total EYFSP score was used by summing the scores across all Early Learning Goals (score range 0-17), where a higher score indicates better school readiness

### Explanatory/Covariates

Ethnicity was coded using 2011 Census categories. IMD 2019 deciles were coded into quintiles based on Lower Super Output Areas. For the EYFSP analyses, child age in months at the time of the EYFSP assessment was used as a continuous variable, calculated using an approximate assessment date of 15 June within the relevant academic year (the EYFSP is completed during the final term and submitted by 30 June) [19].

### Analysis

To describe patterns of sociodemographic characteristics on ASQ-3 and EYFSP outcomes, separate logistic regression analyses were modelled. For the primary analysis, estimating the effect of ASQ-3 on EYFSP, multivariable regression models were estimated including child sex, ethnicity, and IMD quintiles as categorical covariates. The intention of this model is to estimate the effect of ASQ-3 performance on EYFSP outcomes, hence ethnicity, child sex, and IMD quintile were considered confounders.

For all regression models, the binary outcomes of ASQ-3 and EYFSP were used. A sensitivity analysis used multivariate linear regression with continuous ASQ-3 and EYFSP scores.

To investigate if the estimated effect of ASQ-3 on EYFSP varied by sociodemographic characteristics, we estimated three binary logistic models with interactions by child sex, ethnicity, and IMD quintile using a likelihood ratio test to compare the fit of the models with and without the interactions [20].

A descriptive analysis to describe the overlap between achieving / not achieving a GLD on both the ASQ-3 and the EYFSP calculated the proportion of children who did or did not achieve a GLD on the ASQ-3 and on the EYFSP.

### Missing data / incomplete data

Analyses were conducted using complete-case analysis, including only participants with valid and non-missing data for the outcome and all covariates required for each specific model. Complete case analysis provides unbiased results when the chance of being a complete case does not depend on the outcome, after taking covariates into account [21]. The present analysis therefore assumes that having complete data does not depend on a child’s development itself (measured via ASQ-3 and/or EYFSP), after taking into account the included confounder variables (measured via IMD, ethnicity, and child sex). We believe that this assumption serves as a reasonable approximation in this context (see Figure 1).

**Figure 1.**
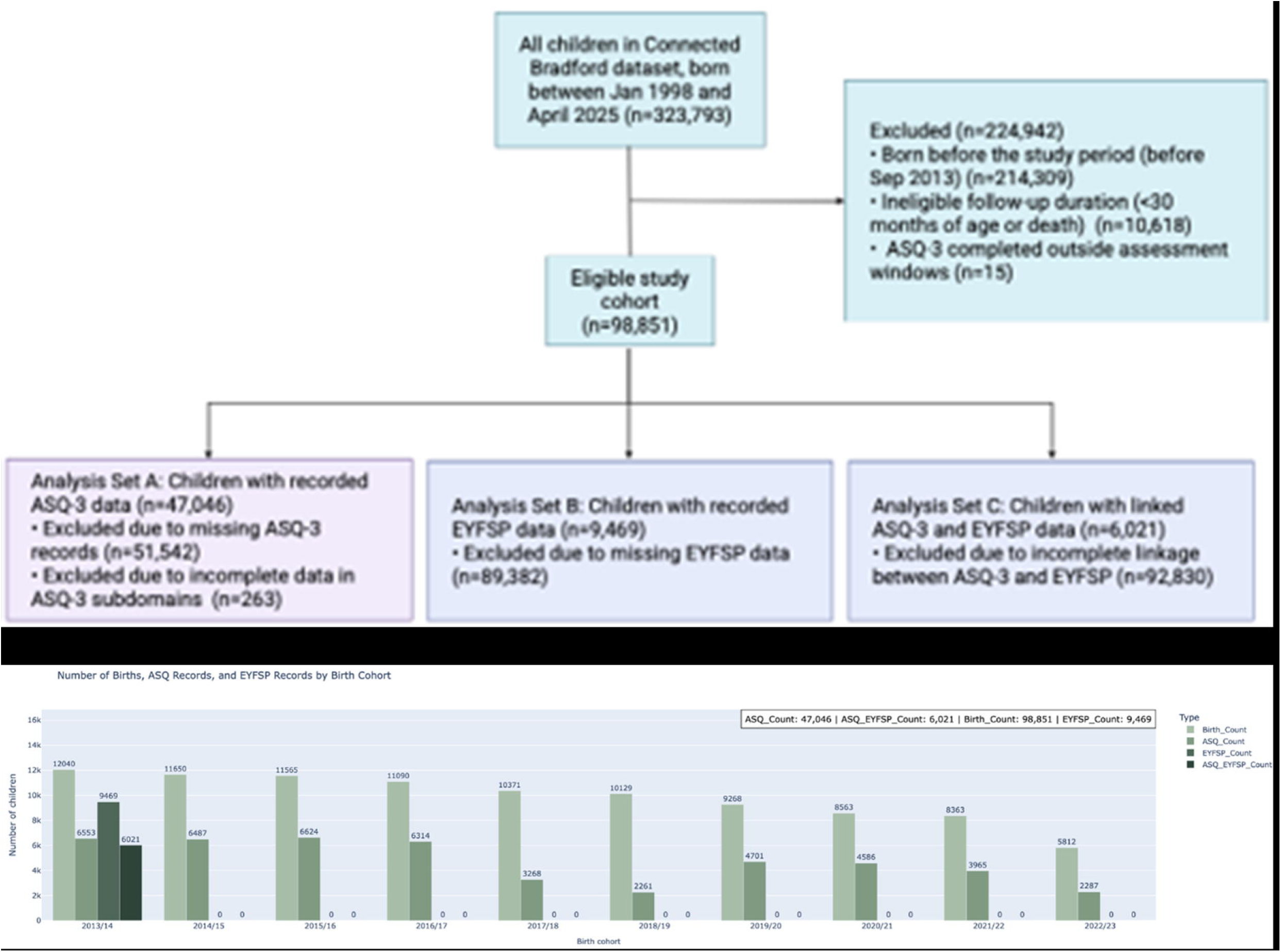
Study eligibility flowchart.

**Figure 2.**
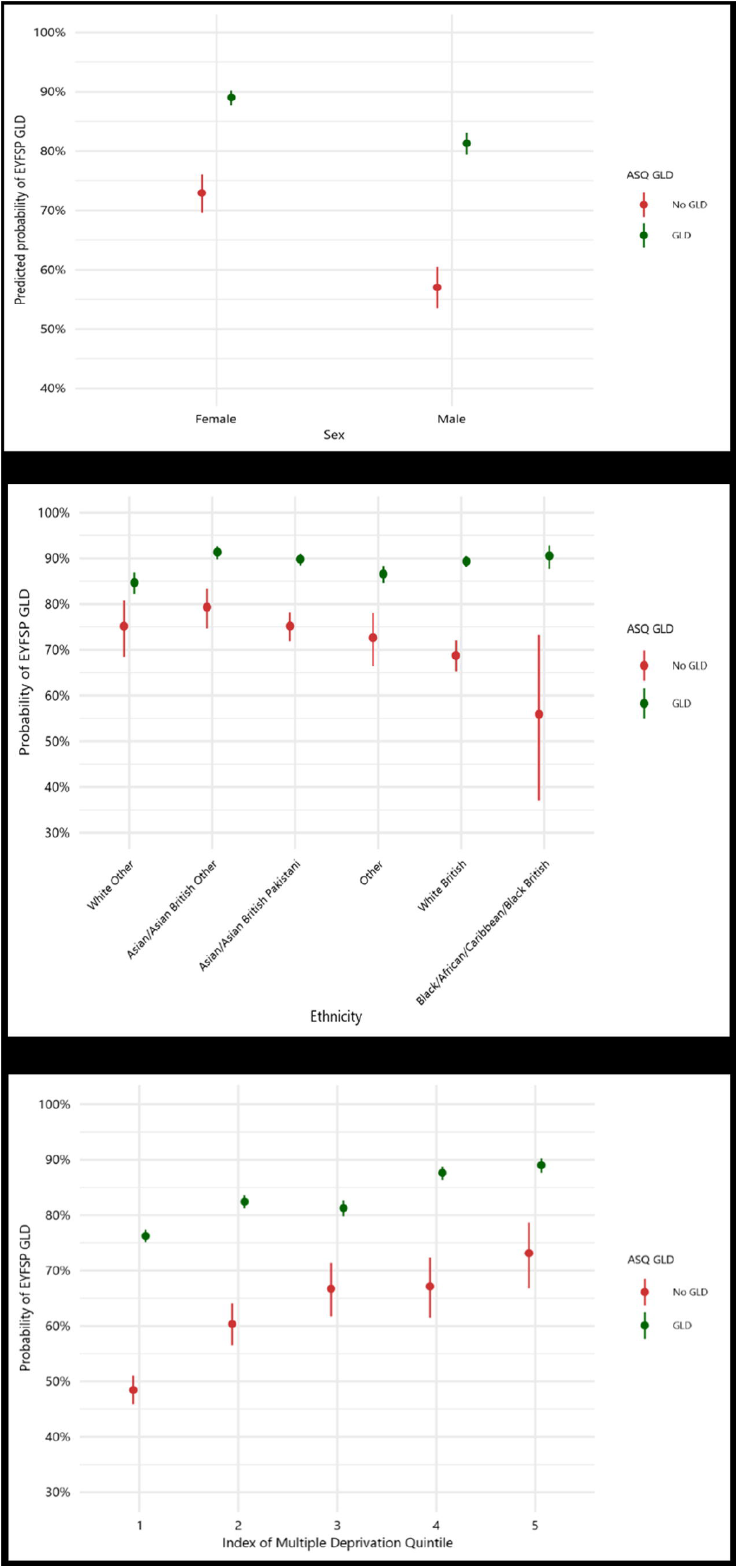
The probability of an EYFSP GLD by ASQ-3 GLD across different (a) ethnic (ordered from smallest to largest difference) and (b) IMD groups (ordered by IMD quintile).

## Results

Figure 1 presents the study flowchart and the available data linkage for each birth year. Of the 98,851 eligible children (born September 2013 to May 2023), 47,309 (48%) had a linked ASQ-3 record and 47,046 had an assessment completed between the ages of 24 and 30 months. At the time of analysis, EYFSP data was only available up to the academic year 2018-2019. Therefore, only children born in 2013-2014 had an EYFSP record (n=9,469), of whom 6,025 had a linked ASQ-3 record.

Table 1 presents the sociodemographic characteristics of the children in the three study samples. The samples have similar proportions of children across key characteristics that are also reflective of the population in the Bradford District [22]: 48-49% female; majority ethnic groups White British (45-49%) and Asian/Asian British Pakistani (30-32%); and 52-55% live in the most deprived IMD quintile.

**Table 1.** Sociodemographic characteristics of children in the ASQ-3 and EYFSP analyses.

| Characteristic | ASQ-3<br>N = 47,046 | EYFSP<br>N = 9,469 | EYFSP + ASQ-3<br>N = 6,021 |
| --- | --- | --- | --- |
| **Sex** |  |  |  |
| Female | 22,854 (48.6%) | 4,529 (47.8%) | 2,921 (48.5%) |
| Male | 24,180 (51.4%) | 4,940 (52.2%) | 3,100 (51.5%) |
| **Ethnicity** |  |  |  |
| Asian/Asian<br>British Other | 2,881 (6.1%) | 619 (6.5%) | 367 (6.1%) |
| Asian/Asian<br>British Pakistani | 15,232 (32.4%) | 2,874 (30.4%) | 1,920 (31.9%) |
| Black/African/Caribbean/Black<br>British | 1,013 (2.2%) | 163 (1.7%) | 70 (1.2%) |
| Other | 3,945 (8.4%) | 1,139 (12.0%) | 458 (7.6%) |
| White<br>British | 21,614 (45.9%) | 4,225 (44.6%) | 2,960 (49.2%) |
| White<br>Other | 2,361 (5.0%) | 449 (4.7%) | 246 (4.1%) |
| **IMD 2019 Quintile** |  |  |  |
| 1 | 22,746 (51.7%) | 4,911 (54.6%) | 3,046 (52.9%) |
| 2 | 8,633 (19.6%) | 1,763 (19.6%) | 1,126 (19.5%) |
| 3 | 4,845 (11.0%) | 931 (10.4%) | 608 (10.6%) |
| 4 | 4,512 (10.3%) | 821 (9.1%) | 570 (9.9%) |
| 5 | 3,270 (7.4%) | 569 (6.3%) | 411 (7.1%) |
| Unknown | 3,040 (6.5%) | 474 (5.0%) | 260 (4.3%) |

The results of the multivariable logistic regression for the study questions are presented in Table 2, and summarised below.

**Table 2.** Multivariable logistic regression of the association between sociodemographic characteristics and achieving a Good Level of Development (GLD) on: ASQ-3; EYFSP; and EYFSP when ASQ-3 GLD is included.

|  | **Outcomes (B, 95% CI)** |  |  |  |  |  |
| --- | --- | --- | --- | --- | --- | --- |
|  | **ASQ-3 odds of achieving GLD\*** |  | **EYFSP odds of achieving GLD\*** |  | **EYFSP odds of achieving GLD by ASQ-3** |  |
|  | **B (95% CI)** | **p** | **B (95% CI)** | **p** | **B (95% CI)** | **p** |
| **ASQ-3 GLD** | NA |  |  |  | 3.18 (2.70 to 3.75) | <0.001 |
| **Age at EYFSP** | NA |  |  |  | 1.17 (1.15 to 1.20) | <0.001 |
| **Gender** |  |  |  |  |  |  |
| Female | Reference |  |  |  |  |  |
| Male | 0.51 (0.48 to 0.53) | <0.001 | 0.56 (0.51 to 0.62) | <0.001 | 0.53 (0.57 to 0.60) | <0.001 |
| **Ethnicity** |  |  |  |  |  |  |
| White British | Reference |  |  |  |  |  |
| Asian/Asian British Other | 0.82 (0.74 to 0.92) | <0.001 | 1.09 (0.89 to 1.33) | 0.411 | 1.34 (1.02 to 1.76) | 0.036 |
| Asian/Asian British Pakistani | 0.80 (0.75 to 0.85) | <0.001 | 1.02 (0.91 to 1.15) | 0.674 | 1.09 (0.94 to 1.27) | 0.240 |
| Black/African/Caribbean/Black British | 0.86 (0.72 to 1.03) | 0.092 | 0.84 (0.59 to 1.19) | 0.309 | 1.05 (0.61 to 1.88) | 0.853 |
| Other | 0.92 (0.84 to 1.02) | 0.110 | 0.84 (0.73 to 0.97) | 0.019 | 0.81 (0.65 to 1.03) | 0.081 |
| White Other | 0.93 (0.83 to 1.06) | 0.272 | 0.52 (0.42 to 0.64) | <0.001 | 0.74 (0.55 to 0.99) | 0.041 |
| **IMD** |  |  |  |  |  |  |
| 5 | Reference |  |  |  |  |  |
| 1 | 0.47 (0.41 to 0.53) | <0.001 | 0.40 (0.32 to 0.50) | <0.001 | 0.39 (0.29 to 0.51) | <0.001 |
| 2 | 0.54 (0.47 to 0.62) | <0.001 | 0.55 (0.43 to 0.69) | <0.001 | 0.58 (0.42 to 0.78) | <0.001 |
| 3 | 0.77 (0.66 to 0.89) | <0.001 | 0.57 (0.44 to 0.74) | <0.001 | 0.56 (0.41 to 0.78) | <0.001 |
| 4 | 0.92 (0.79 to 1.07) | 0.279 | 0.79 (0.60 to 1.03) | <0.078 | 0.85 (0.61 to 1.20) | 0.4 |
| **Number of observations** | **43,996** |  | **8,995** |  | **5761** |  |
| **R<sup>2</sup>** | **0.027** |  | **0.038** |  | **0.118** |  |

\*Note: The sociodemographic differences in the ASQ-3 and EYFSP models represent mutually adjusted model coefficients to describe differences.

Of the 47,046 children with an ASQ-3, 39,187 (83.3%) achieved a GLD. Children less likely to achieve a GLD were: boys (OR 0.51, 95% CI 0.48 to 0.53); children from Asian/Asian British Pakistani, and Asian/Asian British Other ethnicities (0.82, 0.74 to 0.92 and 0.80, 0.75 to 0.85); and children living in the lowest three quintiles of IMD (e.g. lowest quintile: 0.47, 0.41 to 0.53).

Of the 9,469 children with an EYFSP assessment, 6,413 (67.7%) achieved a GLD. Children less likely to achieve a GLD were: Boys (0.56; 0.51 to 0.62); from White Other ethnicity and those in the other ethnic group (0.52, 0.42 to 0.64 and 0.84, 0.73 to 0.97 respectively); and children from the lowest three quintiles of IMD (e.g., lowest quintile 0.40, 0.32 to 0.50).

With sex, ethnicity, age and IMD adjusted, achieving a GLD on the ASQ-3 did estimate the effect of achieving a GLD on the EYFSP - children who achieved a GLD on the ASQ-3 had more than three times the odds of achieving a GLD on the EYFSP (3.18, 2.70 to 3.75).

In the sensitivity analysis using continuous scores the same estimated effect was seen, where an increase of one ASQ-3 goal was estimated an increase of 1.7 in the EYFSP total score (B=1.7, 1.6 to 1.9) (See Supplemental Table S1).

The models with the interaction fitted the data equally well for child sex (x^2^=1.198, p=0.273), whereas the model with the interaction was superior for ethnicity (x^2^=32.617, p<.001) and IMD (x^2^=13.494, p=.009). The estimated effect between ASQ-3 GLD and EYFSP GLD was strongest for Black/African/Caribbean/Black British children, and weakest for White Other children, and was stronger for children living in the lowest IMD quintile.

Table 3 shows high overlap between *not* achieving a GLD on ASQ-3 and EYFSP - children who do not achieve a GLD on the ASQ-3 are likely to also not achieve a GLD on the EYFSP. However, the ASQ-3 misses identification of a high proportion of children (76%) who will also go on to not achieve a GLD on the EYFSP.

**Table 3.** Overlap between GLDs on the ASQ-3 and the EYFSP.

|  | **EYFSP GLD = NO** | **EYFSP GLD = YES** |
| --- | --- | --- |
|  | **N (% overlap)** | **N (% overlap)** |
| **ASQ-3 GLD = NO** | **436 (24%)** | **368 (9%)** |
| **ASQ-3 GLD = YES** | **1392 (76%)** | **3825 (91%)** |
| **Total** | **1828 (100%)** | **4193 (100%)** |

## Discussion

To our knowledge, this is the first study to use linked longitudinal, individual level data to reveal that achieving a GLD on the ASQ-3 provides more than three times the odds of achieving a GLD on the EYFSP. ASQ-3 performance at age two was strongly associated with EYFSP outcomes at age five. The majority of children who do not achieve a GLD on the ASQ-3 do not achieve a GLD on the EYFSP, although many children who later fail to achieve a GLD are not identified by ASQ-3. This reflects the findings from a recent scoping review that suggests that the ASQ-3 is most sensitive when detecting severe delay, with sensitivities for detecting mild-moderate delay being between 23% and 62-77% [23].

In England, the ASQ-3 assessment is currently used as a population level measurement to provide an overview of child development, rather than as an individual screening tool [18,23]. The findings in this study highlights the value of using the ASQ-3 to identify individual children at risk of not achieve a GLD in school and providing evidence-based interventions to address concerns before the school. In order to do so equitably however, there would need to be appropriate interventions available for these children, and equitable access to the two-year review – nationally, 1 in 4 children do not currently receive this review [24].

It is also important to note that these findings highlight that many children already have developmental delay by the age of two. This emphasises the importance of intervening as early as possible, with a focus on the first 1001 days (pregnancy up to the child’s third birthday) which has unequivocal evidence of being the optimal time to prevent lifelong inequality in children’s development and opportunities [25].

This study also highlights previously reported findings that boys and socioeconomically disadvantaged children are less likely to achieve a GLD on the ASQ-3 and EYFSP, but differing patterns for ethnicity were found.

Differences in the strength of the relationship between ASQ-3 and EYFSP by ethnicity and deprivation may reflect both measurement factors and wider social inequities. Whilst the ASQ-3 is self-reported by the child’s parent/carer, the EYFSP is completed by the child’s teacher, meaning that differences in understanding of the questions or concepts, as well as unconscious biases may influence the completion of the assessments by both parents/carers and teachers. There may also be cultural or language differences that make the tools more or less valid for children from some ethnic backgrounds. Whilst the communication and literacy assessments in the ASQ-3 can be answered irrelevant of the child’s dominant language, in the EYFSP language and literacy are based on the child’s English language skills, meaning that children with limited English language ability will do less well on this assessment. Similarly, the ASQ-3 is completed by the child’s parent/carer, the EYFSP is completed by the child’s teacher - differences in understanding of the questions or concepts, as well as unconscious biases may influence completion of both assessments.

Families living in higher socioeconomic circumstances often have access to good quality home learning environments and better-quality early childhood education and care, both of which are known to improve the likelihood of achieving a GLD on the EYFSP [26,27], and these families are more able to actively seek and access additional support services for their children [27-28]. Similarly, families from some ethnic backgrounds can offer their children additional protective factors such as greater social support and/or cohesion that may enhance the likelihood of achieving a GLD on the EYFSP [29, 30].

### Strengths & Limitations

This study reports findings from a single district in England, but includes a sample from multiple ethnic backgrounds and living in areas of high/low deprivation, allowing us to report nuanced, intersectional findings on children at greatest risk of not meeting a GLD on the EYFSP. These are therefore, nationally relevant and should be generalisable to other areas with similar populations, particularly inner-city areas of England. Repeating this analysis in other areas with linked data would allow better understanding of the generalisability.

This study could only link the ASQ-3 to EYFSP scores for children in one academic year (2018-2019). Once education data is available for more academic years it will be important to replicate this analysis to observe if findings remain consistent, particularly given the potential impact of the COVID-19 pandemic on developmental inequities.

The data in CYB could not identify the children living in the district who were eligible, but did not have a two-year ASQ-3 assessment. We were therefore unable to identify if not having a two-year ASQ-3 assessment estimated the effect on EYFSP, an important research question to answer.

We used multivariable models to describe sociodemographic differences in children’s outcomes with adjustment for ethnicity, IMD, and sex simultaneously. These model coefficients therefore should not be interpreted as causal effects, but instead as mutually adjusted descriptions of sociodemographic differences in outcomes. In contrast, our model estimating the impact of ASQ-3 on EYFSP is intended to be interpreted as a causal effect estimate. However, as this study relied on routine data, only a limited number of confounders could be included, which may result in unknown bias in this study. Further analyses with detailed baseline data would further confirm these findings.

## Conclusion

ASQ-3 may be a useful tool to help identify children who would benefit from additional early intervention to help them to achieve a GLD on the EYFSP measure. For children experiencing disadvantage, and for boys, even earlier intervention, from pregnancy onwards, would provide the most effective and equitable ways to enhance child outcomes.

## Data Availability

The data used in this study is a part of the Connected Bradford dataset. Data can be accessed at: https://bradfordresearch.nhs.uk/connected-bradford/governance-and-ethics/

## Data Accessibility

The data used in this study is a part of the CYB dataset. Data can be accessed at: Request for Data Access - Bradford Institute for Health Research. The code for this analysis available in Supplementary Table S2.

## Acknowledgements

The data is provided by the citizens of Bradford and district, and collected by the NHS, DfE and other organisations as part of their care and support. The interpretation and conclusions contained in this study are those of the authors alone. The NHS, DfE and other organisations do not accept responsibility for inferences and conclusions derived from their data by third parties.

## Funding Statement

This is independent research funded by a Wellcome Trust Discovery Award (BiBBS Achieve, 310017/Z/24/Z) and the National Institute for Health and Care Research (NIHR) - YX, RS and HH are supported by NIHR Bradford Health Determinants Research Collaboration (HDRC); JD, MMW & JW are supported by the NIHR Yorkshire and Humber Applied Research Collaboration (ARC-YH; Ref: NIHR200166, see https://www.arc-yh.nihr.ac.uk); JD is further funded by a Population Health Career Scientist Award, NIHR302938 for this research project. The views in this publication are those of the authors and not necessarily those of the NIHR or the Department of Health and Social Care.

## Competing Interests Statement

All authors have completed the ICMJE uniform disclosure form at http://www.icmje.org/disclosure-of-interest/ and declare: no support from any organisation for the submitted work; no financial relationships with any organisations that might have an interest in the submitted work in the previous three years; no other relationships or activities that could appear to have influenced the submitted work.

## Contributorship Statement

JD, DL, KB, PD and KM conceived the idea and designed the study. RS, KS, MMW and JW acquired and managed the clinical data. YX and RS performed the statistical analysis, KM and HH acted as supervisor for all analyses. JD, YX, HH and KM drafted the manuscript, and all authors (JD, YX, RS, HH, DL, KB, PD, KS, MMW, JW and KM) contributed to critical revision of the manuscript for important intellectual content and approved the version to be published. JD is the guarantor and attests that all listed authors meet authorship criteria and that no others meeting the criteria have been omitted

## Notes

### Competing Interest Statement

The authors have declared no competing interest.

### Author Declarations

This study is based on data from Connected Yorkshire which has ethical approval to use data for research purposes (NHS REC 18/YH/0200 & 22/EM/0127).

### Summary of Updates

Following on from reviewers comments, we have amended the paper as follows: We have amended the introduction to add more detail to the previous research literature. We have amended and clarified the methods sections to be clear on the choice of methods and analyses undertaken. We have added further clarity to the sample linkage and to explain why there was large missingness in the EYFSP data. In the Results we added an exploratory analysis of the overlap between ASQ-3 and EYFSP that helps us to better define our recommendations - that is, that children who do not achieve a GLD on the ASQ-3 are likely to not achieve a GLD on the EYFSP, but that the ASQ-3 only identifies 1 in 4 children who will go on to not achieve a GLD on the EYFSP. We have amended the Discussion to be clear about the interpretation of findings

