## Supplemental Table 1 for "Does performance on the Ages and Stages Questionnaire-3 assessment at age 2 estimate the effect on the Early Years Foundation Stage at age 5? A longitudinal observational study using routine data" (from another version of this preprint: version 1, 2026-03-02)

**Supplementary tables**

Table A –Multivariable linear regression of the associations between ASQ-3 total score and sociodemographic characteristics on the outcome of EYFSP total score.

| **Predictors** | **EYFSP total score (0-17)** | |
| --- | --- | --- |
|  | **B (95% CI)** | ***p*** |
| **ASQ-3 composite (0-5)** | 1.7 (1.6 to 1.9) | **<0.001** |
| **Age at EYFSP** | 0.65 | **<0.001** |
| **Gender** |  |  |
| Female |  |  |
| Male | -1.9 (-2.3 to -1.6) | <0.001 |
| **Ethnicity** |  |  |
| White British |  |  |
| Asian/Asian British Other | 0.54 (-0.26 to 1.3) | 0.185 |
| Asian/Asian British Pakistani | 0.14 (-0.31 to 0.59) | 0.546 |
| Black/African/Caribbean/Black British | 0.72 (-1.0 to 2.5) | 0.418 |
| Other | -0.96 (-1.7 to -0.25) | **0.008** |
| White Other | -2.3 (-3.3 to -1.4) | **<0.001** |
| **IMD** |  |  |
| 5 |  |  |
| 1 | -3.8 (-4.6 to -3.1) | **<0.001** |
| 2 | -2.5 (-3.3 to -1.7) | **<0.001** |
| 3 | -2.1 (-3.0 to -1.3) | **<0.001** |
| 4 | -0.50 (-1.4 to 0.38) | 0.262 |
| **Number of observations** | **5761** | |
| **R^2^** | **0.195** | |
